# Stimulation of the Thalamus for Arousal Restoral in Temporal Lobe Epilepsy (START) Clinical Trial: Modulation of Natural Sleep and Focal Seizures

**DOI:** 10.1101/2025.09.18.25335682

**Authors:** Taruna Yadav, Zheng Zhang, Vaclav Kremen, Zan Ahmad, Eva Alden, Joshua P. Aronson, Kimberly Bailey, Jonathan L. Baker, Christopher Benjamin, Benjamin H. Brinkmann, Jordan Burchfield, Krzysztof A. Bujarski, Xi Chen, Eun Young Choi, Kate Christison-Lagay, Devon Cormier, Paul E. Croarkin, Karla Crockett, George Culler, Kristine Dacosta, Eyiyemisi Damisah, Allyson Derry, Catherine Doucet, Eric B. Geller, Jason Gerrard, Joseph Giacino, Nicholas Gregg, Abhijeet Gummadavelli, Jaimie Henderson, Lawrence Hirsch, Jennifer Hong, Matthew Hook, Charlotte A. Jeffreys, Anastasia Kanishcheva, Ruben I. Kuzniecky, Patrice Lauture, Bogdan Patedakis Litvinov, Brian Lundstrom, Stephen Meisenhelter, Steven Messina, Charles Mikell, Sima Mofakham, Grant G. Moncrief, Kyle O’Sullivan, Maxime Oriol, Imran H. Quraishi, Jermaine Robertson, Robert M. Roth, Brian Rutt, Vladimir Sladky, Yinchen Song, Dennis Spencer, George P. Thomas, Jamie Van Gompel, Angela Waszkiewicz, Lydia Wheeler, Courtney Yotter, Christopher R. Butson, Nicholas Schiff, Barbara Jobst, Gregory Worrell, Hal Blumenfeld

## Abstract

Thalamic stimulation has emerged as a promising neuromodulation target for treating disorders of consciousness. Impaired consciousness, a debilitating outcome in temporal lobe epilepsy (TLE) remains a central problem for patients whose seizures cannot be treated pharmacologically and cannot be stopped with conventional surgery or responsive hippocampal stimulation. Although prior studies suggest an essential role of the thalamic intralaminar central lateral (CL) nucleus in arousal and sleep, evidence for a direct effect of thalamic intralaminar stimulation on human arousal has been limited. To address these gaps, the START (stimulation of the thalamus for arousal restoral in TLE) clinical trial investigated the efficacy of bilateral CL thalamic stimulation to restore consciousness during human sleep and TLE seizures. Five patients with medically refractory mesial temporal lobe epilepsy were implanted with an investigational neurostimulator, the Medtronic Summit RC+S^TM^. Optimal CL stimulation parameters were obtained through individualized titration in slow wave sleep, evaluated through analysis of patient movement from video recordings and electrophysiology from simultaneously recorded scalp and hippocampal electroencephalography (EEG). We found that bilateral CL stimulation led to robust arousal from sleep characterized by increased body movements and decreased low frequency power (2-15 Hz) in both cortical and hippocampal EEG during 5 minute epochs of stimulation compared to baseline slow wave sleep. We evaluated impaired consciousness during seizures using verbal and non-verbal behavioral tests administered automatically by smartwatch, and found significant behavioral impairment in three of five patients during seizures with hippocampal stimulation. Administering CL stimulation in patients with impaired consciousness during TLE seizures showed significant improvement in behavioral outcomes in two of three patients, with one patient reaching their baseline performance comparable to non-seizure times. Overall, we found that stimulation of the thalamic CL in TLE patients increased arousal during both sleep and seizures. These findings demonstrate the potential of CL as a therapeutic target for mitigating impaired consciousness in TLE and can serve as a foundation for additional studies to test generalizability of CL stimulation effects on other disorders of consciousness.

## Introduction

Epilepsy affects over 3 million people in the United States (Zack, 2017), with approximately one-sixth unable to achieve seizure control through medications or surgery. Mesial temporal lobe epilepsy (TLE) is the most common form of treatment-resistant epilepsy, comprising 50–60% of patients receiving responsive neurostimulation (Geller et al., 2017; Morrell, 2011). Most individuals with TLE experience disabling focal seizures with impaired consciousness (French et al., 1993), impacting over 200,000 people in the U.S.

Impaired consciousness during and following seizures significantly worsens quality of life, increases the risk of accidents, injury, poor academic and work performance, and social stigma (Charidimou & Selai, 2011; Chen et al., 2014; De Boer, Mula, & Sander, 2008; Nei & Bagla, 2007). Postictal impairment in consciousness may also compromise respiration, contributing to sudden unexpected death in epilepsy (Massey, Sowers, Dlouhy, & Richerson, 2014; Ruthirago, Julayanont, Karukote, Shehabeldin, & Nugent, 2018). Although the primary goal of epilepsy care is to stop seizures, restoring conscious awareness in patients whose seizures cannot be stopped (by medications, surgery or brain stimulation) could significantly improve outcomes. Previous studies suggest potential mechanisms through which TLE seizures cause impaired consciousness through depressed arousal. Intracranial EEG in TLE seizures found strong correlation between impaired consciousness and ictal slow wave activity resembling deep sleep in the extra-temporal brain regions (Blumenfeld, Rivera, et al., 2004; Englot et al., 2010; Yadav et al., 2025). Single photon emission computed tomography (SPECT) imaging reveals reduced blood flow in the frontal and parietal regions, as well as abnormal activity in brainstem and thalamic arousal systems, related to impaired consciousness (Blumenfeld, McNally, et al., 2004; Lee et al., 2002; Rabinowicz, Salas, Beserra, Leiguarda, & Vazquez, 1997). These findings support the “network inhibition hypothesis,” suggesting that limbic seizures inhibit subcortical arousal circuits, leading indirectly to widespread depressed cortical function (Baker et al., 2016; Norden & Blumenfeld, 2002). This hypothesis is further supported by results from animal studies, which have demonstrated that increased activity within subcortical inhibitory regions is strongly associated with depressed subcortical arousal in brainstem and thalamus, especially the thalamic intralaminar central lateral nucleus (CL) (Englot et al., 2008; Englot et al., 2009; Feng et al., 2017; Motelow et al., 2015). Moreover, prior studies in animal models of TLE have shown promise in restoring cortical physiological arousal and behavioral responsiveness during and following limbic seizures through optogenetic or electrical stimulation of subcortical arousal areas, including thalamic CL (Furman et al., 2015; Abhijeet Gummadavelli et al., 2015; Kundishora et al., 2017). These studies therefore suggest that thalamic CL nuclei can be potential neurostimulation target for arousal restoration during TLE seizures.

Disorders of consciousness in other neurological disorders (besides epilepsy) are linked to dysfunction in subcortical–cortical arousal networks (Laureys, Gosseries, & Tononi, 2015; Posner, 2007). Deep brain stimulation (DBS) of the CL nucleus has shown promise in restoring consciousness in patients with chronic disorders (Schiff, 2012; Schiff et al., 2023; Schiff et al., 2007). Similarly, transient impaired consciousness during TLE seizures involves disruption of these same circuits, including CL (Blumenfeld, McNally, et al., 2004; Blumenfeld, Rivera, et al., 2004; Englot et al., 2010; Guye et al., 2006). Studies have shown suppressed CL activity during seizures (Feng et al., 2017; Motelow et al., 2015), and importantly, stimulation of CL in animal models can restore both physiological and behavioral arousal during and after seizures (A. Gummadavelli et al., 2015; Abhijeet Gummadavelli et al., 2015; Kundishora et al., 2017).

Within the domain of sleep research, converging evidence highlights the critical role of CL stimulation in regulating arousal states. Rodent sleep studies have demonstrated that 100 Hz electrical stimulation of the CL produces pronounced effects, including global behavioral arousal, widespread cortical *c*-*fos* activation (Shirvalkar, Seth, Schiff, & Herrera, 2006), sleep-to-wake transitions, and increases in global cortical fMRI signals (Liu et al., 2015). Importantly, translational sleep studies further reveal bilateral depression of CL function during limbic seizures (Feng et al., 2017; Motelow et al., 2015), as well as the capacity to restore both physiological and behavioral arousal via bilateral thalamic CL stimulation during ictal and postictal periods (A. Gummadavelli et al., 2015; Abhijeet Gummadavelli et al., 2015; Kundishora et al., 2017; Xu J, 2017). Complementing these findings, nonhuman primate studies have shown that CL stimulation not only activates intrinsic neuronal populations but also engages *en passant* ascending arousal fibers traversing the intralaminar medial dorsal thalamic tegmental tract (DTTm) (Baker et al., 2016). High-frequency stimulation (100–150 Hz) within this pathway produced rapid and robust improvements in physiological arousal, reaction time sustained attention and working memory capacities in non-human primates (Baker et al., 2016; Shah et al., 2004). In addition, stimulation of the central thalamus has been shown in non-human primates to produce robust physiological and behavioral arousal from sleep and anesthesia (Afrasiabi et al., 2021; Bastos et al., 2021; Redinbaugh et al., 2020; Tasserie et al., 2022).

Collectively, these experimental studies underscore the translational potential of CL stimulation for restoring arousal across species and contexts.

DBS treatment for epilepsy has advanced rapidly, with FDA-approved devices like responsive neurostimulation (RNS, NeuroPace) (Heck et al., 2014; Morrell, 2011) and anterior nucleus thalamic stimulation (Medtronic) (Fisher et al., 2010). Newer investigational systems, such as the Medtronic Summit RC+S^TM^, allow responsive stimulation across multiple brain regions, including novel targets like the CL nucleus. Previous work has also developed the Epilepsy Personal Assistant Device (EPAD), a handheld platform that interfaces with RC+S^TM^ to enable cloud-based data tracking, seizure diaries, and Automatic Responsiveness Testing in Epilepsy (ARTiE) (Kremen et al., 2018; Touloumes et al., 2016; Wheeler et al., 2025). Building on these tools, our goal is to use bilateral CL stimulation via RC+S^TM^ to evaluate efficacy through arousal effects from natural sleep, and to then restore consciousness during TLE seizures unresponsive to conventional RNS, potentially improving quality of life in this population.

## Methods

### Study design

All procedures were in accordance with the institutional review boards for human studies at the three recruitment sites (Yale University School of Medicine, Mayo Clinic and Dartmouth-Hitchcock medical center), as well as National Institutes of Health guidelines. Informed consent was obtained from all patients.

Five patients (P1-P5) with medically refractory temporal lobe epilepsy and mesial temporal onset seizures were enrolled in the study for implantation of an investigational neurostimulator (Medtronic RC+S^TM^) in the hippocampus (HC) and central lateral (CL) intralaminar thalamus.

Figure 1A shows the study design and timeline spanning >=19 months from enrollment. During the baseline phase, patients were asked to maintain a seizure diary and only if the patient reported >=2 disabling seizures (seizures with impaired consciousness) in a month, they were considered eligible for RC+S^TM^ implantation. After implantation and recovery, for the CL stimulus titration phase (Figure 1A), patients were admitted to the inpatient video-EEG monitoring unit with the goal of identifying optimal CL stimulation parameters (current amplitude, frequency, pulse width, electrode contacts and configuration) without side-effects. CL stimulation parameters were titrated per individual patient’s tolerance and tested for their effectiveness in causing arousal from natural sleep. The most effective CL stimulation parameters in sleep titration were used during seizures in the subsequent phases of the study. In the open label hippocampal stimulation phase (OLHC), patients received hippocampal stimulation but no CL stimulation for all detected seizures. In the next, most crucial phase of the study, CL randomization phase, all seizures continued to receive HC stimulation but ∼50% of seizures were randomly assigned to receive additional CL stimulation (stimulated seizures) while other half didn’t receive any CL stimulation (sham seizures). All study personnel including clinicians and patients were blinded to CL stimulation except the device engineers who were not blinded to ensure patient safety. Patients could exit the randomization phase when >= 16 seizures (at least 8 sham and 8 stim seizures) were obtained. In the last phase of the study, open label CL stim + HC stim phase (OLCL), all seizures received both HC and CL stimulation.

**Figure 1.**
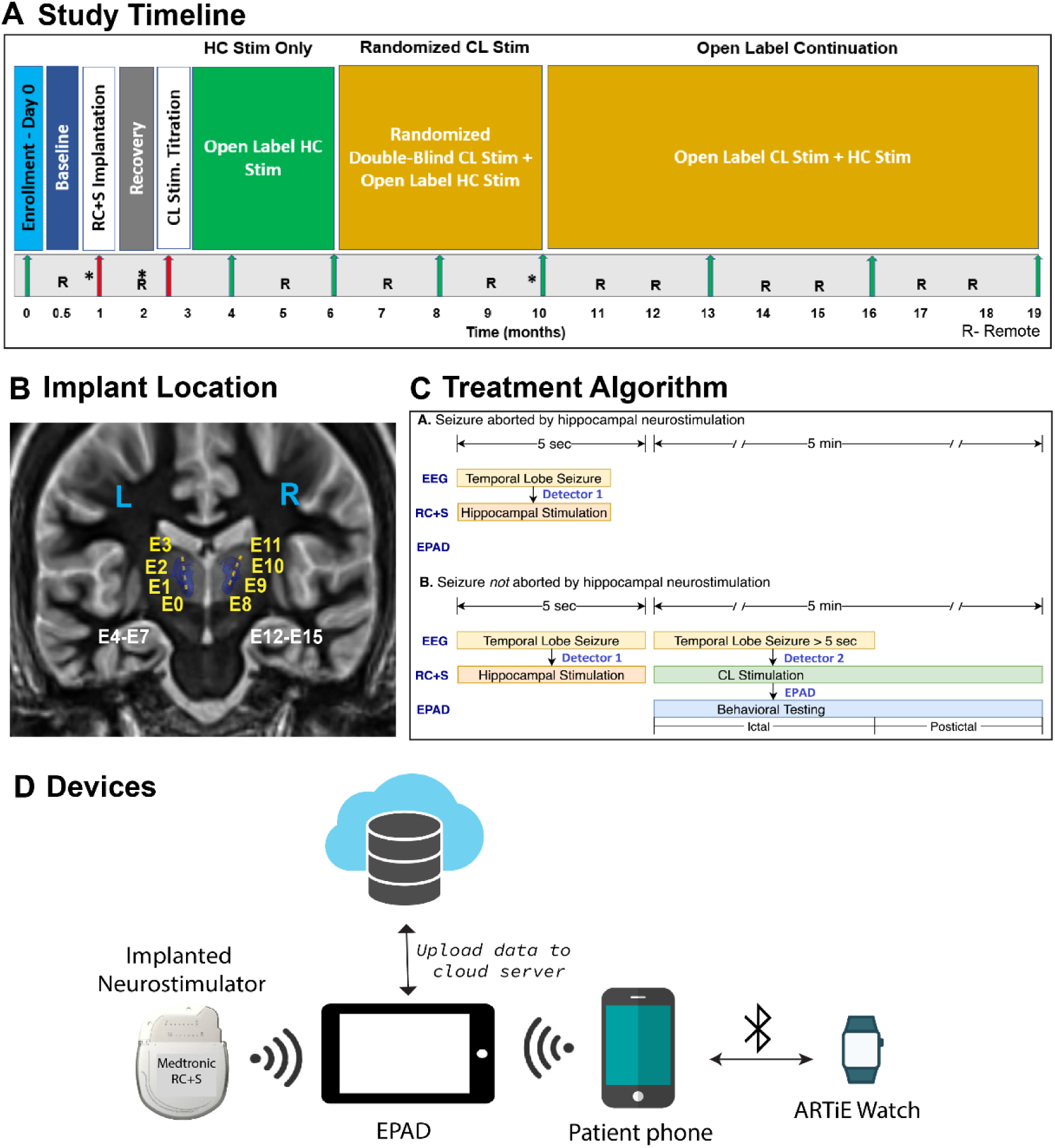
Summary of the START clinical trial design. A) Study timeline shows the different phases each patient went through from enrollment to open label continuation phase spanning >=19 months. Red and green arrows represent in-patient and out-patient visits respectively. * indicates the visits when patient’s eligibility to move to the next study phase was assessed. B) Device implant location depicting the CL targets (in blue), CL electrode contacts (in yellow, left-E0 to E3 and right-E8 to E11) and hippocampal electrode contacts (in white, left E4 to E7 and right-E12 to E15). C) Treatment algorithm summarizes the automatic delivery of HC and CL stimulation and launch of behavioral testing when a seizure is detected. D) Schematic showing communication between different study devices and wireless data upload to server for storage and later processing.

### Electrophysiological data acquisition and processing

All patients were implanted in bilateral HC and bilateral CL except one patient (P4) for whom one lead was placed in the right somatosensory (SS) cortex, one in right HC, and the other two leads in bilateral CL. There were four leads in total for all patients and each lead consisted of four contacts that could be programmed to record or stimulate the brain’s electrical activity (Figure 1B). Intracranial EEG was recorded from four channels at a sampling rate of 500 Hz with 0.85 Hz high-pass analog filter applied. EEG was streamed 24 x 7 from the implanted neurostimulator to the patient tablet (EPAD) and uploaded to cloud server for storage and processing (Figure 1D).

Scalp EEG during the CL inpatient sleep titration phase was recorded at a sampling rate of 256 Hz from 19 standard scalp EEG channels. Bad EEG channels were rejected based on visual review and remaining channels were additionally processed for artifacts due to CL stimulation or movement using the Persyst® proprietary artifact reduction algorithm. Artifact reduced signal was again visually reviewed for any remaining artifacts and removed from the signal. Final cleaned scalp EEG signals were exported and processed in MATLAB 2020a along with intracranial EEG signals. Scalp EEG signal was first high pass filtered at 0.5 Hz (11^th^ order Butterworth filter) and then low pass filtered at 100 Hz (37^th^ order Butterworth filter).

Time-frequency maps were generated in MATLAB using *spectrogram* function with 1-sec non-overlapping Hamming windows at 1 Hz frequency resolution. Spectrograms were generated for each scalp and intracranial EEG channel, normalized at each frequency to the mean power in the baseline period for that frequency and averaged across channels. Normalized power was transformed to dB scale (10*log10) for display.

### Sleep titration video-scalp EEG monitoring and movement scoring

During the sleep titration phase, patients’ video and scalp EEG were recorded overnight alongside intracranial EEG from the neurostimulator. Once the patient was asleep in N2/N3 stage (determined in real time by clinicians based on behavior such as eye closure, no movement and scalp EEG characteristics such as slow-wave sleep, sleep spindles), bilateral CL stimulation was remotely delivered for 5 minutes at previously determined parameters tested in the awake state to ensure no paresthesia or other side effects. CL stimulation was repeated over multiple epochs varying in stimulation current, frequency and pulse width. The effectiveness of CL stimulation in causing arousal from sleep was established based on both behavioral and electrophysiological changes in scalp EEG and HC. Sufficient time (a minimum of approximately 15 minutes) was given between consecutive stimulations to ensure that patients went back to N2/N3 sleep before stimulation was again applied.

For analysis, patient behavior during sleep (5 minutes), arousal (5 minutes) and recovery periods (5 minutes) were observed on video recordings and scored independently by two reviewers. Each 1-sec period was evaluated and given a score of 0 for no movement, score of 1 if it was a simple movement (i.e. one body part or limb moved), score of 2 for a complex movement (i.e. more than one body part/limb moved) and score of 3 for eye opening. For any discrepancies between reviewers, a final score for each 1-sec period was arrived at through consensus between reviewers. Each 1-sec movement period was then assigned a total score = summation of the scores from simple or complex movement and eye movement that ranged from 0 to 5. This way a movement time course was obtained which was smoothened with a 30-sec moving average.

### Treatment algorithm

All patients received responsive HC stimulation at 125 Hz, 90 µs triggered by detection of epileptiform activity on the HC channels. CL stimulation was delivered at frequency of 125 or 40 Hz and pulse width of 90 or 120 µs as identified for each patient during the CL sleep titration phase. Figure 1C summarizes the treatment algorithm. When a seizure was detected by the neurostimulator, responsive bilateral HC stimulation was turned on for 1 second. Seizure status was then continuously checked and if the seizure stopped, HC stimulation was turned off (Figure 1C.a.). If after 5 sec of continuous HC stimulation the seizure did not stop, then HC stimulation was turned off and bilateral CL stimulation was turned on for 5 minutes (Figure 1C.b.). Note that CL stimulation was turned on only for the stimulated (versus sham) seizures in the CL randomization phase, and CL stimulation was turned on for all seizures in the OLCL phase. Seizures during OLHC phase did not receive any CL stimulation.

### Behavioral assessment and analysis

To assess whether CL stimulation was effective in restoring consciousness during seizures, an automatic behavioral testing called Automatic Responsiveness Testing in Epilepsy (ARTiE) (Touloumes et al., 2016; Wheeler et al., 2025) was launched within 5 seconds of the start of CL stimulation on the patient’s smartwatch (ARTiE Watch). The ARTiE behavioral testing was triggered by onboard seizure detection on the RC+S^TM^ device, relayed via the EPAD and smartphone (Figure 1 D). ARTiE testing was triggered regardless of the status of CL stimulation i.e. all seizures in the OLHC, CL randomization and OLCL phases had ARTiE testing if they were not stopped by 5 seconds of continuous HC stimulation. Details of the testing items and scoring method are previously described in (Wheeler et al., 2025). Briefly, the test consisted of 18 verbal and non-verbal test items assessing the patient’s responsiveness and memory functions as summarized in Table S *1*. The entire ARTiE watch testing sequence lasted ∼7 minutes, intended to include ictal and postictal periods. ARTiE watch testing was administered automatically through audio commands and required verbal and non-verbal responses from the patient. Test stimuli and responses were saved as audio recordings on the EPAD and uploaded to the cloud server for later analysis. Two independent reviewers retrospectively evaluated patient responses on each test item and scored them on a scale of 0 (no response) to 3 (correct response) (Table S1). Besides testing during seizures, each patient’s baseline performance was established by conducting two ARTiE tests per week at non-seizures times throughout the course of the study.

## Results

### Robust arousal from sleep with CL stimulation

During the sleep titration study, we evaluated the effectiveness of bilateral CL stimulation in causing arousal from natural sleep. Figure 2 displays a representative snapshot of patient P1’s sleep/awake state and scalp EEG in a 20-sec period. The patient was asleep during the baseline period as verified both behavioral signs (e.g. eyes closed, no body movements) and scalp EEG changes involving slow-wave activity and sleep spindles. After CL stimulation was applied, the patient showed robust signatures of arousal from sleep including eye-opening, sitting up and turning in bed as well as desynchronization of the scalp EEG and disappearance of slow-wave activity.

**Figure 2.**
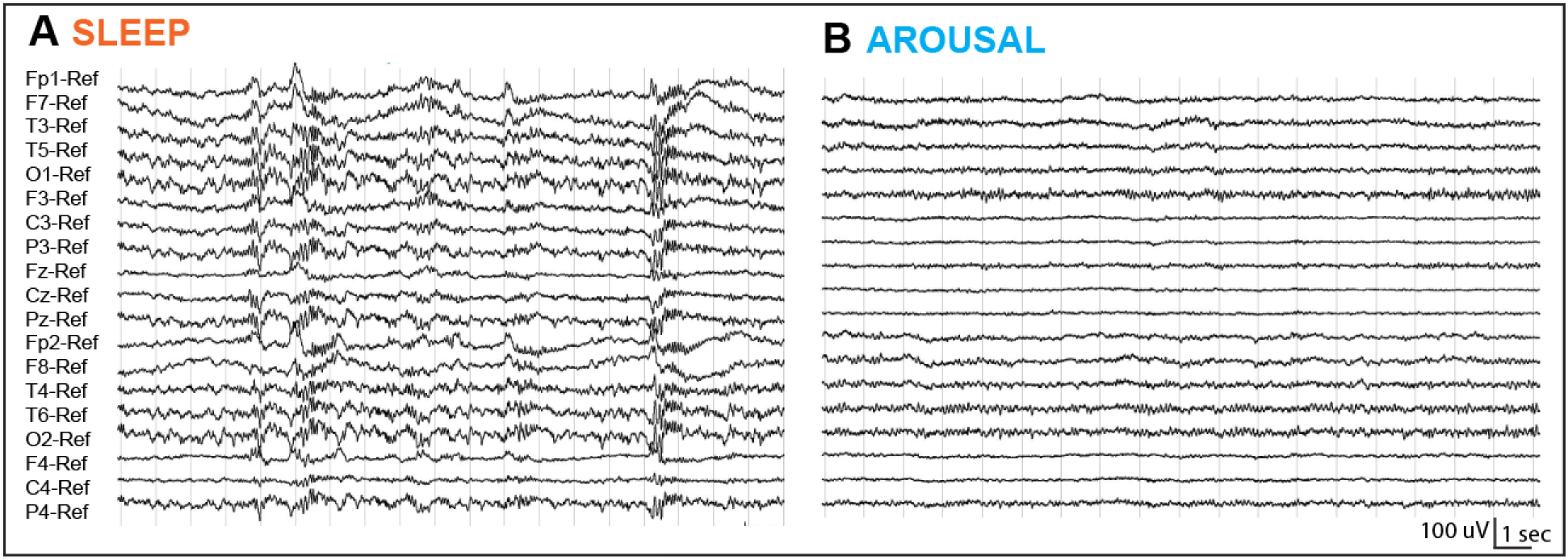
Representative example from overnight scalp EEG recording demonstrating arousal from sleep with CL stimulation for patient P1. Patient scalp EEG (∼20 s), A) natural sleep (prior to CL stimulation) and B) arousal (during CL stimulation).

To quantify arousal from sleep, we computed behavioral and electrophysiological metrics. Figure 3 shows group analysis involving epochs of CL stimulation for each patient that had the most robust arousal from sleep. Figure 3A shows the average time-frequency map across patients for scalp and hippocampal EEG. Data represents normalized power (dB) relative to baseline period (5 min prior to CL stim onset) for the prestimulus baseline, five minutes of stimulation, and five minutes post-stimulus offset. Scalp EEG, left HC and right HC show sustained reduction in low frequency power (2-15 Hz) with onset of CL stimulation compared to baseline power indicating that patients experienced electrophysiological arousal from sleep during the 5-minute stimulation period. EEG power across the scalp and HC remained suppressed for a few minutes after the CL stim was turned off and began to increase towards the end of recovery period indicating patients were slowly going back to sleep.

**Figure 3.**
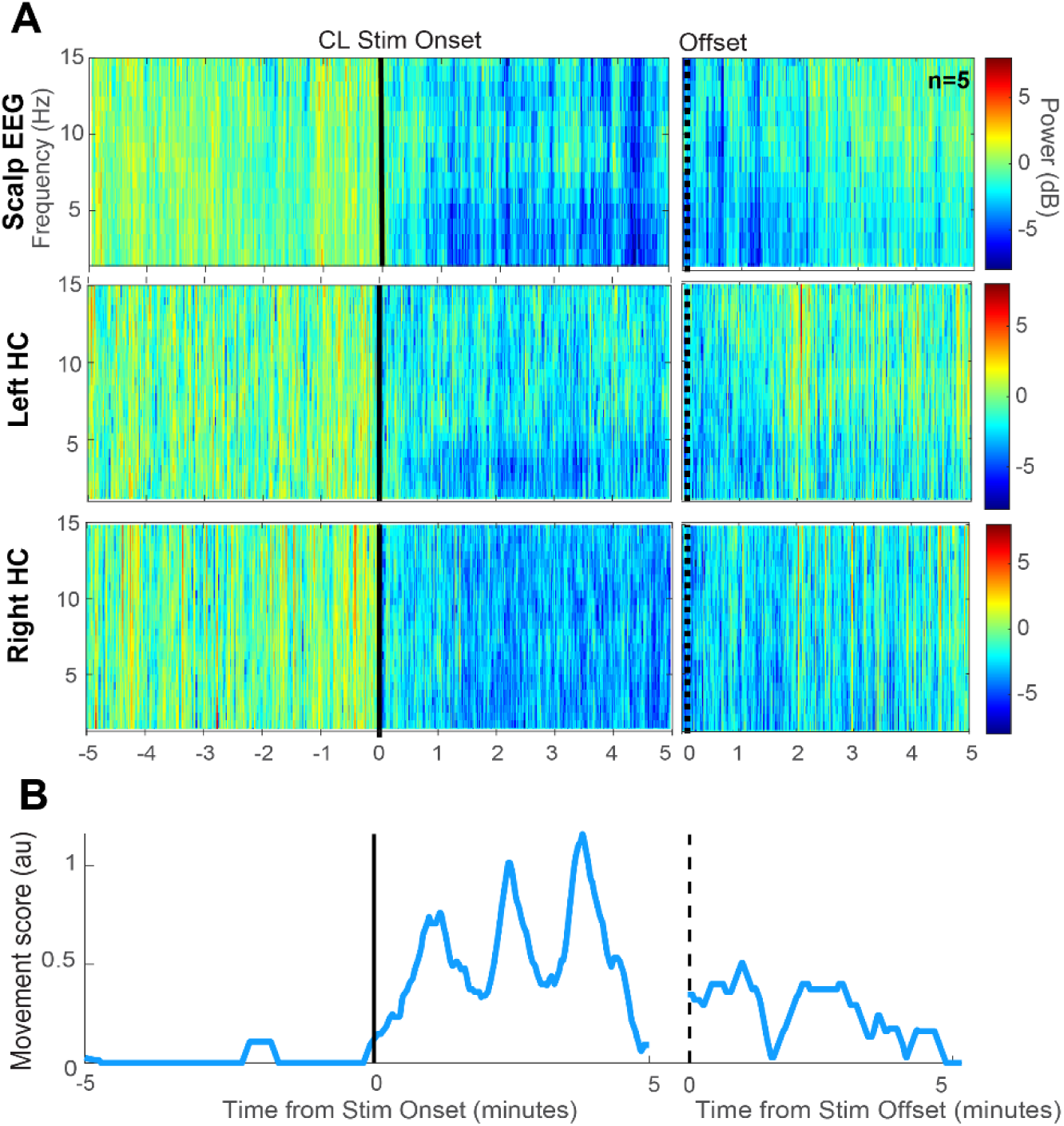
Behavioral and physiological arousal from natural sleep with bilateral CL stimulation. A) Time-frequency maps aligned to CL stimulation onset and offset for scalp EEG, left hippocampus (HC) and right hippocampus. Time-frequency maps were averaged across the CL stim with best arousal from sleep for each patient during 5 minutes of baseline (sleep), 5 minutes of stimulation and 5 minutes of recovery periods immediately after stimulation end. Data show signal power (in log 10 scale) normalized to mean power during baseline period. B) Mean movement score computed across same time periods as A) with best arousal from sleep in each patient.

Figure 3B shows the average movement score across patients during the same baseline, stimulation and recovery periods. As expected, during the baseline period when patients were in stage N2/N3 sleep they had an average movement score close to 0. After the CL stimulation was turned on, movement score was increased throughout stimulation period indicating that patients moved more during this period than baseline. Once the CL stim was turned off (after the dashed black line), patients movement remained somewhat increased compared to baseline, as the patients went back into sleep.

These behavioral and electrophysiological patterns thus validate that bilateral CL stimulation effectively aroused the patients from natural sleep. Effective CL stimulation parameters (corresponding to best arousal) for each patient were next used for stimulation during seizures.

### Primary behavioral outcome of CL stimulation during seizures

We compared ARTiE scores during the baseline (interictal period) and ictal periods to assess the effect of seizures on behavioral performance and whether they caused impaired consciousness. Note that as a condition for enrolment, all patients were required to have at least two seizures per month with impaired consciousness for at least two months prior to device implantation. After device implantation patients began to receive HC stimulation for all seizures during the OLHC phase (Figure 1A). Table 1 summarizes the mean (SEM) interictal and ictal scores during the OLHC and CL randomization phases for the 5 patients (P1-P5). Only those seizures that received hippocampal stimulation but not CL stimulation were included in this analysis, to examine the amount of impairment in each patient during seizures with HC stimulation, but without CL stimulation. Patients P1, P2 and P3 showed significantly lower (p<0.05, paired two-sample t-test) ictal scores compared to interictal scores, during both the OLHC and CL randomization phases suggesting that seizures impaired their behavior even with HC stimulation (Table 1). On the other hand, patients P4 and P5 achieved nearly perfect ictal scores comparable to their interictal values (∼3) and thus did not show significant reduction in behavioral performance during seizures (Table 1). Based on the results of ARTiE testing, we concluded that patients P4 and P5 did not have significant impaired consciousness during seizures with HC stimulation, and because their performance was nearly perfect during seizures, effectiveness of CL stimulation on behavior could not be tested. Therefore, primary outcome measures evaluating the effect of CL stimulation on behavior during seizures were analyzed only for the remaining patients P1, P2 and P3, who had significant behavioral impairment during seizures even with HC stimulation.

**Table 1.** Mean (± SEM) interictal and ictal ARTiE scores during OLHC and CL randomization phase. Ictal scores are from seizures that received HC stimulation but did not receive CL stimulation. P-values were obtained from paired two-sample t-test. Number of seizures used for mean computation are represented by N.

| Patient | OLHC Phase |  |  | Randomization Phase |  |  |
| --- | --- | --- | --- | --- | --- | --- |
|  | Interictal score | Ictal score | P value | Interictal score | Ictal score | P value |
| 1 | 2.993 ± 0.005<br>(N=23) | 2.076 ± 0.072<br>(N=11) | < 0.0001 | 2.985 ± 0.016<br>(N=84) | 2.480 ± 0.084<br>(N=11) | < 0.0001 |
| 2 | 2.975 ± 0.010<br>(N=29) | 1.509 ± 0.122<br>(N=6) | < 0.0001 | 2.992 ± 0.004<br>(N=48) | 1.303 ± 0.223<br>(N=9) | < 0.0001 |
| 3 | 2.998 ± 0.005<br>(N=33) | 1.799 ± 0.155<br>(N=19) | < 0.0001 | 2.991 ± 0.005<br>(N=47) | 2.192 ± 0.153<br>(N=13) | < 0.0001 |
| 4 | 2.953 ± 0.030<br>(N=26) | 2.944 ± 0.056<br>(N=1) | 0.765 | 2.915 ± 0.028<br>(N=62) | 2.920 ± 0.043<br>(N=16) | 0.822 |
| 5 | 2.985 ± 0.006<br>(N=22) | 2.806 ± 0.099<br>(N=10) | 0.083 | 2.975 ± 0.007<br>(N=194) | 2.852 ± 0.068<br>(N=12) | 0.065 |

Figure 4 A and C show the results of the CL randomization phase comparing ARTiE scores for sham vs. stimulated seizures for patients P1, P2 and P3. Figure 4 B and D show similar comparisons of ARTiE scores from seizures without CL stimulation (i.e. from OLHC phase) and seizures with CL stimulation (i.e. from OLCL phase). Time-course data represent mean (and SEM) ARTiE score across seizures for each test instruction in sequence. Bar plots represent mean and SEM scores across all test instructions. Time course depicts how severely patient’s consciousness was affected by the seizure. Initially, ARTiE scores for both types of seizures (sham/OLHC and stim/OLCL) are low at the beginning of the seizure and as the patient recovers from the seizure (during later test instructions), ARTiE scores gradually increase towards baseline scores except poor scores in the “memory2” test instruction. Low memory scores (“memory1”) indicates that patients were severely impaired when the two words were asked to be repeated and memorized for later. Low “memory 2” scores suggest memory deficits i.e. patient didn’t remember the words that were given at the beginning of ARTiE testing.

**Figure 4.**
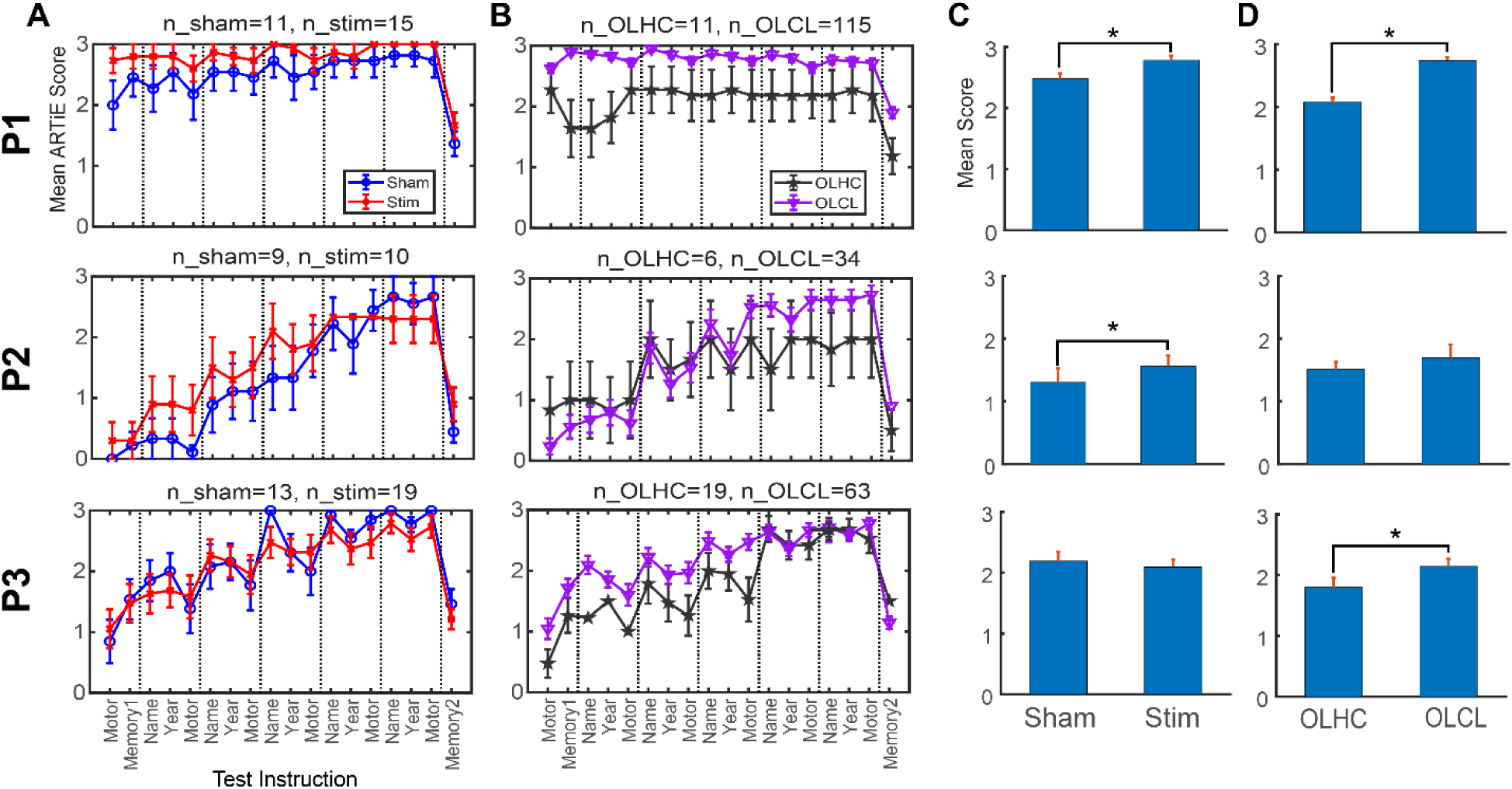
Time course (mean and SEM) showing behavioral performance in ARTiE testing of 3 patients (P1, P2 and P3) comparing seizures without and with CL stimulation during A) CL randomization phase and B) OLHC and OLCL phases. C) and D) correspond to respective overall mean (SEM) scores across seizures. * represents significant difference between scores (p<0.05, paired two sample t-test).

The primary outcome of CL stimulation on recovery from impaired consciousness can be clearly seen for patients P1 and P2. These patients performed better on ARTiE testing during CL stimulated seizures compared to sham. Both these patients showed significant improvement in ARTiE scores (p<0.05, two-sample t-test) when stimulated in the central lateral thalamus with patient P1 remarkably scoring nearly as well as their baseline score (∼3). We similarly compared performance during seizures from the OLHC and OLCL phases and found that mean ARTiE scores were higher for seizures receiving CL stimulation and reached significance (p<0.05) in two of three patients (Figure 4 D). Overall, our results demonstrate that stimulating the CL during seizures led to improved behavioral scores in two of three patients compared to when CL was not stimulated.

## Discussion

The START clinical trial examined the efficacy and safety of bilateral thalamic CL stimulation to restore consciousness during seizures in TLE patients. The main results demonstrated that CL stimulation produced robust and reproducible arousal from natural sleep, validating the titrated stimulation parameters as effective in activating arousal networks in humans. Second, in two of the three patients who experienced impaired consciousness during seizures despite HC stimulation, CL stimulation improved scores on an automatic behavioral testing tool (ARTiE). These results demonstrate, for the first time in humans, that direct stimulation of the CL nucleus can mitigate seizure-related disorders of consciousness.

### Restoral of Arousal during Seizures

The principal clinical finding of this study is the enhancement of arousal and behavioral responsiveness during TLE seizures through therapeutic stimulation of the thalamic CL nucleus. In two of the three patients who exhibited impaired consciousness, ARTiE scores were significantly elevated during CL stimulation compared to sham stimulation. These findings are consistent with the network inhibition hypothesis, which proposes that TLE seizures disrupt subcortical arousal systems, resulting in widespread cortical depression and consequent loss of consciousness (Blumenfeld, 2012; Norden & Blumenfeld, 2002). Complementary animal research has further shown that seizures are associated with suppressed activity in the thalamic CL nucleus, and that thalamic stimulation can restore behavioral responsiveness (Feng et al., 2017; Abhijeet Gummadavelli et al., 2015; Motelow et al., 2015). Together, the present study provides the first direct evidence in humans that stimulation of the thalamic CL nucleus can improve seizure-induced suppression of arousal networks, thereby restoring the capacity for behavioral responsiveness. Notably, CL stimulation in our trial was designed to engage arousal circuits directly, shifting the therapeutic emphasis from seizure elimination alone to the broader goal of preserving consciousness and enhancing quality of life. This conceptual realignment parallels recent advances in the treatment of disorders of consciousness, where CL stimulation has been shown to promote wakefulness and purposeful behavior (Schiff, 2012; Schiff et al., 2023; Schiff et al., 2007).

The clinical implications of these findings are substantial. Restoring consciousness during seizures has the potential to markedly enhance patient safety, autonomy, and overall quality of life. Impaired consciousness remains one of the most disabling aspects of epilepsy, associated with accidents, driving restrictions, unemployment, and social stigma (Chen et al., 2014; De Boer et al., 2008; Nei & Bagla, 2007). If neuromodulation can restore responsiveness during seizures, patients may retain greater independence, reduce injury risk, and experience improved psychosocial functioning. Moreover, restoration of responsiveness may mitigate postictal respiratory compromise and reduce the risk of sudden unexpected death in epilepsy (SUDEP) (Massey et al., 2014; Ruthirago et al., 2018).

Two patients (P4, P5) demonstrated near-maximal ARTiE scores regardless of CL stimulation. One possible explanation is that HC stimulation may have attenuated seizure severity or altered limbic–thalamocortical network dynamics, thereby improving arousal and behavioral performance during seizures independently of CL input. These findings, despite the small sample size, demonstrate the possible efficacy of HC stimulation alone in improving behavioral responsiveness during seizures and should be investigated further. Rigorous behavioral testing during seizures, as was done in the present study with the ARTiE watch, can facilitate future studies of the effects of HC stimulation on behavior in TLE. In addition, such testing could be useful for identifying ideal candidates for CL stimulation in future studies. Implementing these methodological refinements will be pivotal for optimizing patient selection and enhancing therapeutic precision in future interventions aimed at restoring consciousness via the thalamic CL stimulation.

### Robust Arousal from Sleep

The observation that bilateral CL stimulation consistently produced arousal from N2/N3 sleep is an important translational validation of preclinical findings. Rodent studies demonstrated that 100 Hz stimulation of the CL induces widespread cortical *c*-*fos* activation, sleep-to-wake or anesthesia-to-wake transitions, and enhanced cortical fMRI signals (Abhijeet Gummadavelli et al., 2015; Kundishora et al., 2017; Liu et al., 2015; Shirvalkar et al., 2006). Similarly, in nonhuman primates, stimulation of the CL and its *en passant* fibers within the intralaminar medial dorsal thalamic tegmental tract (DTTm) resulted in rapid improvements in arousal, reaction time, sustained attention, and working memory (Baker et al., 2016; Shah et al., 2004), as well as arousal from sleep and anesthesia (Afrasiabi et al., 2021; Bastos et al., 2021; Redinbaugh et al., 2020; Tasserie et al., 2022). Our results extend these findings to humans with epilepsy, confirming that CL stimulation exerts powerful effects on global arousal networks, producing robust behavioral and electrophysiological arousal from natural sleep.

Our findings, significantly reduced delta and power and increased body movements with CL stimulation, are consistent with evidence that thalamic circuits can actively shift cortical activity into a desynchronized, aroused mode. This suggests that while the electrophysiological markers of arousal may differ across species and brain regions, the thalamic CL represents a conserved pathway through which wakefulness can be restored across contexts, including sleep and seizures.

### Limitations and Future Directions

The small sample size (n = 5) limits statistical power and generalizability. Moreover, only three patients demonstrated ictal impairment of consciousness, and stimulation efficacy could only be meaningfully assessed in these patients. Follow-up studies with larger patient population will be needed to validate these findings.

Second, parameter titration was individualized during sleep arousal testing. While effective, this approach introduces variability that may complicate replication. Systematic exploration of stimulation frequency, amplitude, and pulse width is needed to establish standardized protocols.

Third, although ARTiE testing offers a rigorous and reproducible measure of responsiveness, it primarily captures overt behavioral performance. Consciousness is multidimensional, encompassing subjective experience and covert awareness, which may not be fully assessed by ARTiE. Incorporating multimodal measures, including fMRI, EEG connectivity, or pupillometry, could provide convergent evidence of consciousness restoration.

Future studies should prioritize larger randomized controlled trials to confirm the efficacy and long-term safety of thalamic CL stimulation. Closed-loop stimulation offers a promising next step. Platforms such as RC+S^TM^ can detect seizures in real time and deliver stimulation only when impairment occurs, potentially improving responsiveness while reducing unnecessary stimulation.

## Data Availability

All data generated in the present study will be made available from the corresponding author upon reasonable request.

## Acknowledgements

We thank Abbey Becker, Thaddeus Brink, Jon Giftakis, Benjamin Isaacson, David Linde and Robert Raike at Medtronic for their technical support in this study. We thank Tyler Hamilton for data management; Starr Guzman, Irina Korytov and Kathy Polak for administrative support; Samuel Heck, Amy Hummel and Zi Kai (Kyle) Soo for their assistance with regulatory processes; Manichanh Howell for data entry; and Delana Weis for efforts in documentation. This work was supported by NIH/NINDS UG3/UH3 NS112826.

## Supplementary information

**Table S1:**
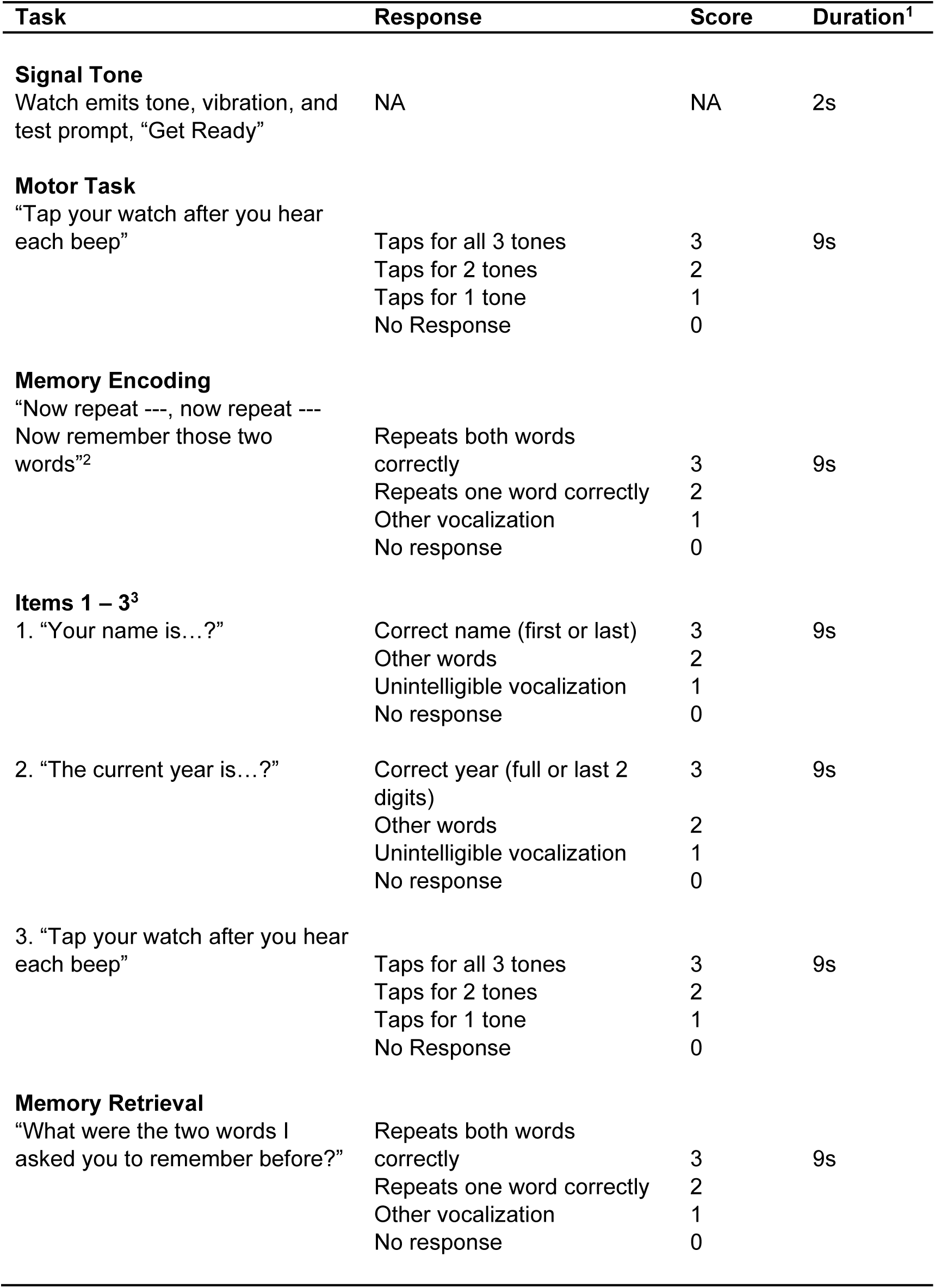
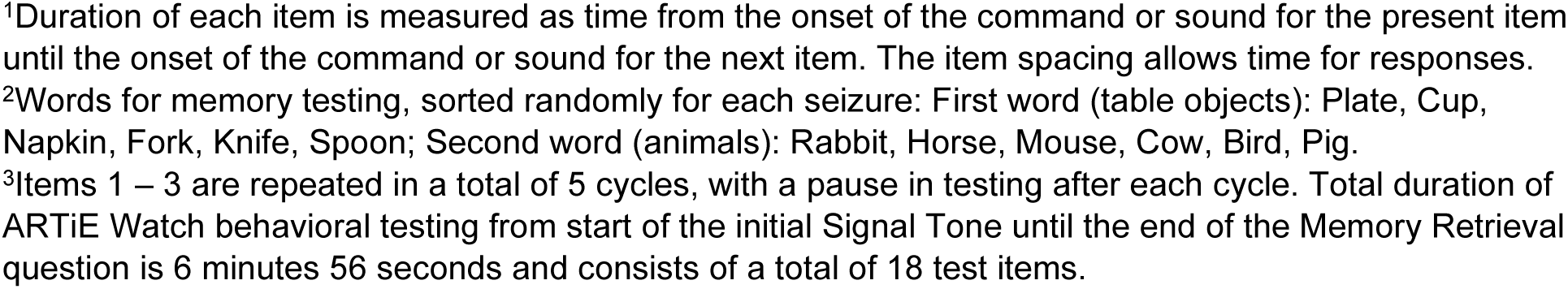
Automatic Behavioral Testing in Epilepsy (ARTiE) Watch Behavioral Testing Protocol (reusing with permission from (Wheeler et al., 2025))

| Task | Response | Score | Duration<sup>1</sup> |
| --- | --- | --- | --- |
| **Signal Tone** |  |  |  |
| Watch emits tone, vibration, and test prompt, "Get Ready" | NA | NA | 2s |
| **Motor Task** |  |  |  |
| "Tap your watch after you hear each beep" | Taps for all 3 tones | 3 | 9s |
|  | Taps for 2 tones | 2 |  |
|  | Taps for 1 tone | 1 |  |
|  | No Response | 0 |  |
| **Memory Encoding** |  |  |  |
| "Now repeat ---, now repeat ---<br>Now remember those two words"<sup>2</sup> | Repeats both words correctly | 3 | 9s |
|  | Repeats one word correctly | 2 |  |
|  | Other vocalization | 1 |  |
|  | No response | 0 |  |
| **Items 1 – 3<sup>3</sup>** |  |  |  |
| 1. "Your name is...?" | Correct name (first or last) | 3 | 9s |
|  | Other words | 2 |  |
|  | Unintelligible vocalization | 1 |  |
|  | No response | 0 |  |
| 2. "The current year is...?" | Correct year (full or last 2 digits) | 3 | 9s |
|  | Other words | 2 |  |
|  | Unintelligible vocalization | 1 |  |
|  | No response | 0 |  |
| 3. "Tap your watch after you hear each beep" | Taps for all 3 tones | 3 | 9s |
|  | Taps for 2 tones | 2 |  |
|  | Taps for 1 tone | 1 |  |
|  | No Response | 0 |  |
| **Memory Retrieval** |  |  |  |
| "What were the two words I asked you to remember before?" | Repeats both words correctly | 3 | 9s |
|  | Repeats one word correctly | 2 |  |
|  | Other vocalization | 1 |  |
|  | No response | 0 |  |

<sup>1</sup>Duration of each item is measured as time from the onset of the command or sound for the present item until the onset of the command or sound for the next item. The item spacing allows time for responses.
<sup>2</sup>Words for memory testing, sorted randomly for each seizure: First word (table objects): Plate, Cup, Napkin, Fork, Knife, Spoon; Second word (animals): Rabbit, Horse, Mouse, Cow, Bird, Pig.
<sup>3</sup>Items 1 – 3 are repeated in a total of 5 cycles, with a pause in testing after each cycle. Total duration of ARTiE Watch behavioral testing from start of the initial Signal Tone until the end of the Memory Retrieval question is 6 minutes 56 seconds and consists of a total of 18 test items.

## Notes

### Competing Interest Statement

The authors have declared no competing interest.

### Clinical Trial

NCT04897776

### Author Declarations

Ethics committee/IRB of Yale University School of Medicine, Mayo Clinic, and Dartmouth-Hitchcock medical center gave ethical approval for this work.

## References

Afrasiabi, M., Redinbaugh, M. J., Phillips, J. M., Kambi, N. A., Mohanta, S., Raz, A., … Saalmann, Y. B. (2021). Consciousness depends on integration between parietal cortex, striatum, and thalamus. Cell systems, 12(4), 363–373.

Baker, J. L., Ryou, J.-W., Wei, X. F., Butson, C. R., Schiff, N. D., & Purpura, K. P. (2016). Robust modulation of arousal regulation, performance, and frontostriatal activity through central thalamic deep brain stimulation in healthy nonhuman primates. Journal of neurophysiology.

Bastos, A. M., Donoghue, J. A., Brincat, S. L., Mahnke, M., Yanar, J., Correa, J., … Brown, E. N. (2021). Neural effects of propofol-induced unconsciousness and its reversal using thalamic stimulation. elife, 10, e60824.

Blumenfeld, H. (2012). Impaired consciousness in epilepsy. The Lancet Neurology, 11(9), 814–826.

Blumenfeld, H., McNally, K. A., Vanderhill, S. D., Paige, A. L., Chung, R., Davis, K., … Novotny, E. J. (2004). Positive and negative network correlations in temporal lobe epilepsy. Cerebral cortex, 14(8), 892–902.

Blumenfeld, H., Rivera, M., McNally, K. A., Davis, K., Spencer, D. D., & Spencer, S. S. (2004). Ictal neocortical slowing in temporal lobe epilepsy. Neurology, 63(6), 1015–1021.

Charidimou, A., & Selai, C. (2011). The effect of alterations in consciousness on quality of life (QoL) in epilepsy: searching for evidence. Behavioural neurology, 24(1), 83–93.

Chen, W. C., Chen, E. Y., Gebre, R. Z., Johnson, M. R., Li, N., Vitkovskiy, P., & Blumenfeld, H. (2014). Epilepsy and driving: potential impact of transient impaired consciousness. Epilepsy & Behavior, 30, 50–57.

De Boer, H. M., Mula, M., & Sander, J. W. (2008). The global burden and stigma of epilepsy. Epilepsy & Behavior, 12(4), 540–546.

Englot, D. J., Mishra, A. M., Mansuripur, P. K., Herman, P., Hyder, F., & Blumenfeld, H. (2008). Remote effects of focal hippocampal seizures on the rat neocortex. Journal of Neuroscience, 28(36), 9066–9081.

Englot, D. J., Modi, B., Mishra, A. M., DeSalvo, M., Hyder, F., & Blumenfeld, H. (2009). Cortical deactivation induced by subcortical network dysfunction in limbic seizures. Journal of Neuroscience, 29(41), 13006–13018.

Englot, D. J., Yang, L. I., Hamid, H., Danielson, N., Bai, X., Marfeo, A., … Motelow, J. E. (2010). Impaired consciousness in temporal lobe seizures: role of cortical slow activity. Brain, 133(12), 3764–3777.

Feng, L., Motelow, J. E., Ma, C., Biche, W., McCafferty, C., Smith, N., … Xiao, B. (2017). Seizures and sleep in the thalamus: focal limbic seizures show divergent activity patterns in different thalamic nuclei. Journal of Neuroscience, 37(47), 11441–11454.

Fisher, R., Salanova, V., Witt, T., Worth, R., Henry, T., Gross, R., … Labar, D. (2010). Electrical stimulation of the anterior nucleus of thalamus for treatment of refractory epilepsy. Epilepsia, 51(5), 899–908.

French, J. A., Williamson, P. D., Thadani, V. M., Darcey, T. M., Mattson, R. H., Spencer, S. S., & Spencer, D. D. (1993). Characteristics of medial temporal lobe epilepsy: I. Results of history and physical examination. Annals of Neurology: Official Journal of the American Neurological Association and the Child Neurology Society, 34(6), 774–780.

Furman, M., Zhan, Q., McCafferty, C., Lerner, B. A., Motelow, J. E., Meng, J., … Deisseroth, K. (2015). Optogenetic stimulation of cholinergic brainstem neurons during focal limbic seizures: Effects on cortical physiology. Epilepsia, 56(12), e198–e202.

Geller, E. B., Skarpaas, T. L., Gross, R. E., Goodman, R. R., Barkley, G. L., Bazil, C. W., … Cole, A. J. (2017). Brain-responsive neurostimulation in patients with medically intractable mesial temporal lobe epilepsy. Epilepsia, 58(6), 994–1004.

Gummadavelli, A., Kundishora, A. J., Willie, J. T., Andrews, J. P., Gerrard, J. L., Spencer, D. D., & Blumenfeld, H. (2015). Improving level of consciousness in epilepsy with neurostimulation. Neurosurg Focus, 38(6), E10.

Gummadavelli, A., Motelow, J. E., Smith, N., Zhan, Q., Schiff, N. D., & Blumenfeld, H. (2015). Thalamic stimulation to improve level of consciousness after seizures: evaluation of electrophysiology and behavior. Epilepsia, 56(1), 114–124.

Guye, M., Régis, J., Tamura, M., Wendling, F., Gonigal, A. M., Chauvel, P., & Bartolomei, F. (2006). The role of corticothalamic coupling in human temporal lobe epilepsy. Brain, 129(7), 1917–1928.

Heck, C. N., King-Stephens, D., Massey, A. D., Nair, D. R., Jobst, B. C., Barkley, G. L., … Gwinn, R. P. (2014). Two-year seizure reduction in adults with medically intractable partial onset epilepsy treated with responsive neurostimulation: final results of the RNS System Pivotal trial. Epilepsia, 55(3), 432–441.

Kremen, V., Brinkmann, B. H., Kim, I., Guragain, H., Nasseri, M., Magee, A. L., … Nelson, N. (2018). Integrating brain implants with local and distributed computing devices: a next generation epilepsy management system. IEEE journal of translational engineering in health and medicine, 6, 1–12.

Kundishora, A. J., Gummadavelli, A., Ma, C., Liu, M., McCafferty, C., Schiff, N. D., … Blumenfeld, H. (2017). Restoring conscious arousal during focal limbic seizures with deep brain stimulation. Cerebral cortex, 27(3), 1964–1975.

Laureys, S., Gosseries, O., & Tononi, G. (2015). The neurology of consciousness: *cognitive neuroscience and neuropathology*: Academic Press.

Lee, K. H., Meador, K. J., Park, Y. D., King, D. W., Murro, A. M., Pillai, J. J., & Kaminski, R. J. (2002). Pathophysiology of altered consciousness during seizures: subtraction SPECT study. Neurology, 59(6), 841–846.

Liu, J., Lee, H. J., Weitz, A. J., Fang, Z., Lin, P., Choy, M., … Mitra, P. (2015). Frequency-selective control of cortical and subcortical networks by central thalamus. elife, 4, e09215.

Massey, C. A., Sowers, L. P., Dlouhy, B. J., & Richerson, G. B. (2014). Mechanisms of sudden unexpected death in epilepsy: the pathway to prevention. Nature Reviews Neurology, 10(5), 271–282.

Morrell, M. J. (2011). Responsive cortical stimulation for the treatment of medically intractable partial epilepsy. Neurology, 77(13), 1295–1304.

Motelow, J. E., Li, W., Zhan, Q., Mishra, A. M., Sachdev, R. N. S., Liu, G., … Chu, V. (2015). Decreased subcortical cholinergic arousal in focal seizures. Neuron, 85(3), 561–572.

Nei, M., & Bagla, R. (2007). Seizure-related injury and death. Current neurology and neuroscience reports, 7(4), 335–341.

Norden, A. D., & Blumenfeld, H. (2002). The role of subcortical structures in human epilepsy. Epilepsy & Behavior, 3(3), 219–231.

Posner, J. B. (2007). Plum and Posner’s diagnosis of stupor and coma (Vol. 71): OUP USA.

Rabinowicz, A. L., Salas, E., Beserra, F., Leiguarda, R. C., & Vazquez, S. E. (1997). Changes in regional cerebral blood flow beyond the temporal lobe in unilateral temporal lobe epilepsy. Epilepsia, 38(9), 1011–1014.

Redinbaugh, M. J., Phillips, J. M., Kambi, N. A., Mohanta, S., Andryk, S., Dooley, G. L., … Saalmann, Y. B. (2020). Thalamus modulates consciousness via layer-specific control of cortex. Neuron, 106(1), 66–75.

Ruthirago, D., Julayanont, P., Karukote, A., Shehabeldin, M., & Nugent, K. (2018). Sudden unexpected death in epilepsy: ongoing challenges in finding mechanisms and prevention. International Journal of Neuroscience, 128(11), 1052–1060.

Schiff, N. D. (2012). Moving toward a generalizable application of central thalamic deep brain stimulation for support of forebrain arousal regulation in the severely injured brain. Annals of the New York Academy of Sciences, 1265(1), 56–68.

Schiff, N. D., Giacino, J. T., Butson, C. R., Choi, E. Y., Baker, J. L., O’Sullivan, K. P., … Chua, J. (2023). Thalamic deep brain stimulation in traumatic brain injury: a phase 1, randomized feasibility study. Nature medicine, 29(12), 3162–3174.

Schiff, N. D., Giacino, J. T., Kalmar, K., Victor, J. D., Baker, K., Gerber, M., … Kobylarz, E. J. (2007). Behavioural improvements with thalamic stimulation after severe traumatic brain injury. Nature, 448(7153), 600–603.

Shah, A. S., Bressler, S. L., Knuth, K. H., Ding, M., Mehta, A. D., Ulbert, I., & Schroeder, C. E. (2004). Neural dynamics and the fundamental mechanisms of event-related brain potentials. Cerebral cortex, 14(5), 476–483.

Shirvalkar, P., Seth, M., Schiff, N. D., & Herrera, D. G. (2006). Cognitive enhancement with central thalamic electrical stimulation. Proceedings of the National Academy of Sciences, 103(45), 17007–17012.

Tasserie, J., Uhrig, L., Sitt, J. D., Manasova, D., Dupont, M., Dehaene, S., & Jarraya, B. (2022). Deep brain stimulation of the thalamus restores signatures of consciousness in a nonhuman primate model. Science advances, 8(11), eabl5547.

Touloumes, G., Morse, E., Chen, W. C., Gober, L., Dente, J., Lilenbaum, R., … Si, Y. (2016). Human bedside evaluation versus automatic responsiveness testing in epilepsy (ARTiE). Epilepsia, 57(1), e28–e32.

Wheeler, L., Kremen, V., Mersereau, C., Ornelas, G., Yadav, T., Cormier, D., … Sladky, V. (2025). Automatic responsiveness testing in epilepsy with wearable technology: The ARTiE Watch. Epilepsia, 66(1), 104–116.

Xu J, G. M., Pok JY, McCafferty CP, Feng L, Gummadavelli A, Kundishora AJ, Gerrard JL, Laubach M, Schiff ND, Blumenfeld H. (2017). Behavioral assessment of intralaminar thalamic neurostimulation to improve consciousness during the postictal period of seizures. Paper presented at the Society for Neuroscience. http://www.abstractsonline.com/pp8/index.html#!/4376/presentation/10291

Yadav, T., Litvinov, B. P., Culler, G. W., Kumar, A., Khalaf, A., Song, Y., … Blumenfeld, H. (2025). Low and high frequency signatures of impaired consciousness in temporal lobe seizures. bioRxiv, 2025.2007.2001.662627. doi:10.1101/2025.07.01.662627

Zack, M. M. (2017). National and state estimates of the numbers of adults and children with active epilepsy—United States, 2015. MMWR. Morbidity and mortality weekly report, 66.

